# Assessment of knowledge, Attitudes and Practices regarding Hepatitis B virus among Nurses in Bangladesh: a cross-sectional study

**DOI:** 10.1101/2024.05.12.24307258

**Authors:** Salekur Rahman, Sadhan Kumar Das, Zaki Farhana, Md Abu Bakkar Siddik, Anjan Kumar Roy, Piue Dey, Shuvojit Kumar Kundu, Md Anwar Hossain, S M Shahinul Islam, Anton Abdulbasah Kamil, Jahan Ara Khanam, Mohammad Meshbahur Rahman

## Abstract

**Background:** Hepatitis B virus (HBV) infection is a worldwide issue. Nurses are particularly at risk of occupational HBV contamination. In Bangladesh, there is little understanding of the knowledge, attitude, and practices (KAP) of nurses regaarding HBV. Therefore, the study aimed to assess the KAP of nurses regarding HBV.

**Methods:** This cross-sectional study was conducted among 120 nurses through face-to-face interviews through a semi-structured questionnaire using a convenient sample technique. Different statistical tools including frequency distribution, Pearson Chi-square test, and t-tests were used in data analysis.

**Results:** The average age of the participants was 34.5 years where majority of them were 31-40 years old (50.0%) and female (83%). This study revealed that 25.0% had good knowledge, 43.3% exhibited a good (positive) attitude, and 56.7% of nurses demonstrated good practices regarding HBV. Conversely, nurses exhibit a poor knowledge rate of 30.0%, a poor attitude rate of 30.8%, and a poor practice rate of 29.2% regarding HBV. The independent sample t-test and one-way ANOVA demonstrated that nurses’ from rural residences had a significantly higher knowledge [0.80 (0.79-0.82) versus 0.78 (0.77-0.79)]; p=0.001] regarding HBV. Similarly, female [mean attitude score: 0.88 (0.85-0.91) versus 0.77 (0.86-0.69); p=0.009] and nurses from Muslim [0.79(0.61-0.97) versus 0.79(0.76-0.83); p=0.035] faiths has significantly higher positive attitude and good practice behaviors compared to their counterparts.

**Conclusions:** Increasing the vaccination coverage rate of all nurses, as well as implementing additional techniques for preventing exposure in the workplace, training programs on HBV infection, including PEP, comes highly recommended.

## Introduction

Infectious diseases, those caused by the hepatitis B virus (HBV) are among the most significant threats to public health across the globe. Recent estimates suggest that around one-third of the total population of the globe has been infected with HBV [1]. Globally, HBV poses a serious threat to public health as a silent killer [2]. According to the information provided by the World Health Organization (WHO), estimates that 240 million people (or one in thirty) globally suffer from a chronic HBV infection, making up roughly one-third of all people who have ever been infected with the virus [2]. Global viral hepatitis is a serious public health issue that affects people everywhere. Hepatocellular carcinoma, cirrhosis, chronic liver disease, and even fulminant hepatitis can result from this inflammation of the liver [3]. Globally, the prevalence of HBV Surface Antigen which indicates the presence of infection, varies substantially depending on geographic location and vaccination efficacy. It ranges from 3% to 6% [4,5]. Over 2 billion individuals globally exhibit signs of current or previous HBV infection, and 350 million are chronic HBV carriers. There are an estimated 80 million HBV carriers in the Southeast Asian region, or roughly 6% of the overall population [6].

Healthcare workers (HCWs), particularly nurses, are very susceptible to contracting HBV in healthcare environments. The incidence rate of HBV among HCWs is about 2 to 10 times greater than that of the general population worldwide [7,8]. Contact with non-intact skin, insufficiently sterilized medical equipment, and percutaneous or mucous exposure to infectious blood or bodily fluids were indicators of risk for HBV infection in the setting of healthcare workers [9]. The typical risk of developing HBV infection from intravascular insertion to contaminated blood is analyzed to be between 6% and 30%, whereas the risk for the virus that causes human immunodeficiency virus is around 0.3% [10].

HCWs including nurses in underdeveloped nations have a significant risk of being exposed to HBV due to the high prevalence of the virus in the general population and inadequate healthcare facilities [11–13]. One example is cross-sectional research that was carried out in Ethiopia. The results of this study revealed, 7.3% of HCWs were infected by HBV, but just 0.9% of non-HCWs who took part in the survey were found to be infected [12]. The greater risk of HBV contamination amongst HCWs in non-developed countries might be because of the widespread casual handling of contaminated materials, reuse of improperly sanitized medical instruments, and an unadvanced waste disposal system [14–16]. As part of the same healthcare delivery system as HCWs, healthcare trainees have the same occupational risk of HBV. Lack of expertise, training, duty stress, and exhaustion may increase trainees’ risk of inadvertent exposure [17,18].

Adherence to universal measures, such as the usage of barrier protection like hand gloves, thorough sterilization of medical equipment, well-managed hospital waste procedures, and immunization, may effectively prevent HBV infection [19,20]. Additionally, prevention upon exposure may be used as a method of preventing HBV following unintentional contact with infected blood or bodily fluids [21,22]. Concerning HBV infection, however, studies have shown that there is a significant knowledge gap among healthcare workers, particularly nurses.

According to the findings of research conducted in Vietnam, for instance, 74.4% of medical students exhibited a lack of awareness of the routes of the transmission of HBV and their perception of risk [23]. In Cambodia, it was discovered that there is a lack of understanding among public sector HCWs on the transmission route, vaccination, diagnosis, and treatment of HBV [24]. According to the results of research conducted in Sudan, approximately fifty percent of the HCWs had insufficient information regarding HBV, and more than 50% of the midwives and nurses did not finish the immunization schedule for HBV [25]. The healthcare personnel in India, on the other hand, have enough information and protocols regarding HBV infection, and they have gotten an adequate number of immunizations [26].

Bangladesh is going through an epidemiological transition with significant reductions in fatality due to acute, infectious, and parasitic diseases and increases in non-communicable, degenerative, and chronic diseases over the last 20 years [27–33]. There is currently a significant research gap in Bangladesh regarding the knowledge, attitude, and practice (KAP) of HBV, particularly from the perspective of nurses. Several studies have been undertaken among the general population, medical students, and barbers, all of which reached the same conclusion: most participants had insufficient KAP about HBV [34–36]. It is essential to have appropriate KAP to avoid infection among healthcare workers, especially nurses, who are the front-line workers. Hence, this study aimed to cover this research gap. The sole objective of this study was to evaluate the KAP related to HBV among nurses working at Rajshahi Medical College Hospital, a tertiary hospital in Bangladesh. Additionally, the study intended to address the existing research gap and provide valuable insights to policymakers for the development of appropriate policies.

## Materials and Methods

### Ethical consent and permission for data collection

The permission of this study was approved by the Institutional Animal, Medical Ethics, Biosafety and Biosecuirity Committee (IAMEBBC) of Institute of Biological Sciences, University of Rajshahi **[Ref. Memo No. 12(22)/320/IAMEBBC/IBSc]**. Both written and verbal consent was taken from each participant before initiating the interview for data collection. A brief introduction on the aims and objectives of the study was given first and then, the written consent translated in Bengali. Participants who were agreed with the consent were finally included in the study.

### Study setting, sample size, and participants

This study was a cross-sectional study conducted covering all the administrative wards of Rajshahi Medical College Hospital in Bangladesh from 1st January to August December 2023.

A convenient sampling technique was followed for this study. The sample size of the study was calculated by using the below formula:

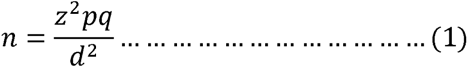

Where,

n= assumed/ desired sample size
z= the standard normal deviation 1.96;
p= percentage of knowledge is 91.5%= 0.915
q= 1-p
d= Degree of error (5%) = 0.05

During the literature search, the proportion of knowledge of good knowledge was high (91.5%) was p=0. 915 [37].

Using the equation (1), the sample size for study when p=0.915 is

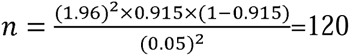

### Inclusion and exclusion criteria

The inclusion criteria of this study were: □) nurses of Rajshahi Medical College hospital; □) nurses aged more than 18 years old; iii) nurses who were Bangladeshi by birth; iv) respondents who provided consent to participate. The exclusion criteria for the study were: i) nurses from the other hospitals visiting RMCH during the study period; ii) nurses holding dual citizenship; iii) nurses who worked at RMCH but were admitted to the hospital due to illness; iv) who did not provide consent to participate.

### Outcome measures

#### Sociodemographic measures

Age (Below 30 years, 31 to 40 years, Above 41 years), Educational status (Up to Primary, Up to Secondary, Up to Higher Secondary, Graduation and above), Monthly family income in Taka (Below 30000 Taka, 30001 to 40000 Taka, Above 40001 Taka), Highest nursing degree (Diploma in Nursing, BSc in Nursing, Master of Science in Nursing, MPH), Area of residence (Urban, Rural), Religion (Muslim, Non-Muslim), Job title/designation of the respondents (Staff Nurse, Senior Staff Nurse, Nursing In-charge), Marital status (Ever Married, Never Married), Work area (Medicine, Surgery, ICU, CCU) were taken as sociodemographical and related measures for this current study.

#### Knowledge, attitude, and practice measurement

Different kinds of questionnaires were used by the researchers in the earlier studies to examine KAP about HBV [34,35]. In the current study, to assess the KAP among the nurses we used a questionnaire containing 36 questions and three sub-sections on knowledge about HBV, attitude towards HBV, and practices regarding HBV. The questionnaire’s reliability and consistency, as shown by a Cronbach’s alpha of 0.87, seem to be flawless for the research.

#### Data analysis

The study has used several statistical descriptive tools and techniques to analyze data. Using Microsoft Excel, the data was compiled. Data was modified, cleaned, and prepared before analysis was undertaken using SPSS 26.0 software. The dataset’s multicollinearity was examined using the correlation matrix. With the use of the Kolmogorov-Smirnov test, we were able to confirm that the data was independent, not dependent, and that there was no multicollinearity. The socioeconomic status differences were shown using a frequency table. We used the Pearson chi-square test to find the factors that were significantly related to KAP. Later ANOVA and t-tests were performed.

To complete the knowledge, attitude, and practice score, we used 16 variables related to Hepatitis B. The average scoring technique was used in computation and descriptive statistics including percentile was observed. According to the percentile approach, the knowledge was classified into three levels: poor knowledge (<25% Percentile, Cut-point: ≥0.7273); Average knowledge (26%-74% Percentile, Cut-point: 0.7273-0.8408); and good knowledge (75% and above percentile, Cut-point: ≤ 0.76).

In attitude score, we used 7 variables related to Hepatitis B. According to the percentile approach, the attitude was classified into three levels: poor attitude (<25% Percentile, Cut-point: ≥0.75); Average attitude (26%-74% Percentile, Cut-point: 0.76-0.875); and good attitude (75% and above percentile, Cut-point: ≤ 0.876).

In the practice score, we used 11 variables related to Hepatitis B. According to the percentile approach, the attitude was classified into three levels: poor practice (<25% Percentile, Cut-point: ≥0.625); Average practice (26%-74% Percentile, Cut-point: 0.626-0.874); and good practice (75% and above percentile, Cut-point: ≤ 0875).

## Results

### Socio-demographic characteristics of the respondents

Table 1 shows the socio-demographic characteristics of the respondents. It was found that the majority of the respondents (50.0%) were aged between 31 and 40 years followed by 17.5% of respondents aged 41 to highest. About 70.8% had completed graduation while 37.5% had completed a Diploma in nursing professional education. Almost half of the participants had a monthly family income of 30001-40000 BDT. Urban participants were more prevalent since the study covered participants from the Rajshahi Medical College where most of them 97.5% were Muslim.

**Table 1.** Socio-demographic characteristics of respondents.

| **Characteristics** | **Frequency**<br>**f(%)** | **Percent**<br>**f(%)** |
| --- | --- | --- |
| **Age** |  |  |
| Below 30 years | 39 | 32.5 |
| 31 to 40 years | 60 | 50.0 |
| Above 41 years | 21 | 17.5 |
| **Sex of the respondents** |  |  |
| Male | 21 | 18 |
| Female | 99 | 83 |
| **Educational status** |  |  |
| Up to Primary | 1 | 0.8 |
| Up to Secondary | 7 | 5.8 |
| Up to Higher Secondary | 27 | 22.5 |
| Graduation and above | 85 | 70.8 |
| **Monthly family income in Taka** |  |  |
| Below 30000 Taka | 44 | 36.7 |
| 30001 to 40000 Taka | 57 | 47.5 |
| Above 40001 Taka | 19 | 15.8 |
| **Highest nursing degree** |  |  |
| Diploma in Nursing | 45 | 37.5 |
| BSc in Nursing | 32 | 26.7 |
| Mater of Science in Nursing | 5 | 4.2 |
| MPH | 38 | 31.7 |
| **Area of residence** |  |  |
| Urban | 106 | 88.3 |
| Rural | 14 | 11.7 |
| **Religion of the respondents** |  |  |
| Muslim | 117 | 97.5 |
| Non-Muslim | 3 | 2.5 |
| **Job title/designation of the respondents** |  |  |
| Staff Nurse | 4 | 3.3 |
| Senior Staff Nurse | 108 | 90.0 |
| Nursing In-charge | 8 | 6.7 |
| **Marital status** |  |  |
| Ever Married | 107 | 89.2 |
| Never Married | 13 | 10.8 |
| **Work area** |  |  |
| Medicine | 32 | 26.7 |
| Surgery | 29 | 24.2 |
| ICU | 17 | 14.2 |
| CCU | 5 | 4.2 |

### Knowledge regarding Hepatitis B Virus among respondents

Table 2 represents participants’ knowledge about HBV. Among the 120 respondents, the majority of the respondents 97.5% knew hepatitis, 96.7% heard of a disease termed HBV, 99.2% knew hepatitis is a viral disease, 95.0% agreed that HBV affects liver disease, 94.2% agreed that HBV causes liver cancer, 85.0% knew HBV affect any age group, 67.5% knew about early symptoms of hepatitis, 95.0% respondents believed HBV affect liver function, 84.2% knew about common symptoms of HBV, 95.8% agreed with HBV be transmitted by un-sterilized syringes, needles, and surgical instruments, 98.3% knew about HBV be transmitted by contaminated blood and blood products, 85.8% knew about HBV transmitted by using blades of the barber/ear and nose piercing, 94.2% agrred HBV transmitted by unsafe sex, 95.8% participants knew about HBV transmitted from mother to child, 42.5% agreed with HBV transmitted by contaminated water/food prepared by a person suffering from these infections, 84.2% agreed with HBV treatable, 39.2% knew about HBV be self-cured by the body, 91.7% knew about vaccination available for HBV, 86.7% have knowledge about the schedule of HBV, 90.8% realized to diagnosis of HBV, 95.0% confirmed to vaccine prevent HBV and 70.0% disagree with HBV affected person can donate bloodm (Table 2).

**Table 2.** Knowledge regarding Hepatitis B Virus among respondents.

| **Knowledge related characteristics** | **No**<br>**f(%)** | **Yes**<br>**f(%)** |
| --- | --- | --- |
| Have heard of Hepatitis | 3 (2.5) | 117 (97.5) |
| Have heard of Hepatitis B | 4 (3.3) | 116 (96.7) |
| Is Hepatitis B being a viral disease | 1 (0.8) | 119 (99.2) |
| Can Hepatitis B affect liver function | 6 (5.0) | 114 (95.0) |
| Can Hepatitis B cause liver cancer? | 7 (5.8) | 113 (94.2) |
| Can Hepatitis B affect any age group? | 18 (15.0) | 102 (85.0) |
| Are the early symptoms of hepatitis B are same as cold and flu? | 39 (32.5) | 81 (67.5) |
| Jaundice is one of the common symptoms of hepatitis B. | 6 (5.0) | 114 (95.0) |
| Are nausea, vomiting, and loss of appetite common symptoms of hepatitis B? | 19 (15.8) | 101 (84.2) |
| Can hepatitis B be transmitted by un-sterilized syringes, needles, and surgical instruments? | 5 (4.2) | 115 (95.8) |
| Can hepatitis B be transmitted by contaminated blood and blood products? | 2 (1.7) | 118 (98.3) |
| Can hepatitis B be transmitted by using blades of the barber/ear and nose piercing? | 17 (14.2) | 103 (85.8) |
| Can hepatitis B be transmitted by unsafe sex? | 7 (5.8) | 113 (94.2) |
| Can hepatitis B be transmitted from mother to child? | 5 (4.2) | 115 (95.8) |
| Can hepatitis B be transmitted by contaminated water/food prepared by a person suffering from these infections? | 69 (57.5) | 51 (42.5) |
| Is hepatitis B treatable? | 19 (15.8) | 101 (84.2) |
| Can hepatitis B be self-cured by the body? | 73 (60.8) | 47 (39.2) |
| Is vaccination available for hepatitis B? | 10 (8.3) | 110 (91.7) |
| If yes, then do you know about the vaccination schedule for both infants & adults? | 16 (13.3) | 16 (86.7) |
| Can a person donate blood if He/she has hepatitis B? | 84 (70.0) | 36 (30.0) |
| Do you know how is hepatitis B diagnosed? | 11 (9.2) | 109 (90.8) |
| Can Hepatitis B be prevented by vaccination? | 6 (5.0) | 114 (95.0) |

### Attitudes regarding Hepatitis B among respondents

Regarding Attitude, three fourth of the respondents 72.5% had a good attitude about HBV as a public problem in Bangladesh, 96.7% had a good attitude about it is important for healthcare professionals, including nurses, to be educated about HBV, 85.0% had a good attitude about HBV is a serious health issue in our community, 86.7% had a good attitude about regular HBV screenings to ensure your safety and the safety of your patients, majority 78.3% good attitude about ever encountered a patient with HBV in your nursing practice, 85.0% attitude feel confident in providing care to the patient with HBV, 91.7% had good attitude about aware of the existing HBV vaccination policy in our healthcare facility and 92.5% had attitude of necessary standard protocols for biomedical waste management (Table 3).

**Table 3.** Distribution of Attitude of Hepatitis B Virus among nurses.

| **Attitude related characteristics** | **No**<br>**f(%)** | **Yes**<br>**f(%)** |
| --- | --- | --- |
| Hepatitis B as a public problem in Bangladesh. | 33 (27.5) | 87 (72.5) |
| Do you think it is important for healthcare professionals, including nurses, to be educated about Hepatitis B? | 4 (3.3) | 116 (96.7) |
| Do you think Hepatitis B is a serious health issue in our community? | 18 (15.0) | 102 (85.0) |
| Are you willing to undergo regular Hepatitis B screenings to ensure your safety and the safety of your patients? | 16 (13.3) | 104 (86.7) |
| Have you ever encountered a patient with Hepatitis B in your nursing practice? | 26 (21.7) | 94 (78.3) |
| If yes, did you feel confident in providing care to the patient with Hepatitis B? | 18 (15.0) | 102 (85.0) |
| Are you aware of the existing Hepatitis B vaccination policy in our healthcare facility? | 10 (8.3) | 110 (91.7) |
| Need on necessary standard protocols for biomedical waste management? | 9 (7.5) | 111 (92.5) |

### Information related to Nurse’s practice regarding Hepatitis B Virus

Table 4 shows the distribution of practice regarding Hepatitis B among Nurses. Among 120 nurses, the majority 91.7% practice receiving HBV vaccination, 91.7% practice to aware of the available treatments for HBV, 60.0% had poor practices about receiving specific training on HBV prevention and management in the last year, 89.2% good practices in preventing HBV transmission in your workplace, 91.7% agreed with practices to educate your patients about the spread and severity of HBV, 70.8% practice to a received vaccine for HBV, 81.7% had good practice to test blood for HBV and 96.7% practice to use sterile instruments, gloves, gowns.

**Table 4:**
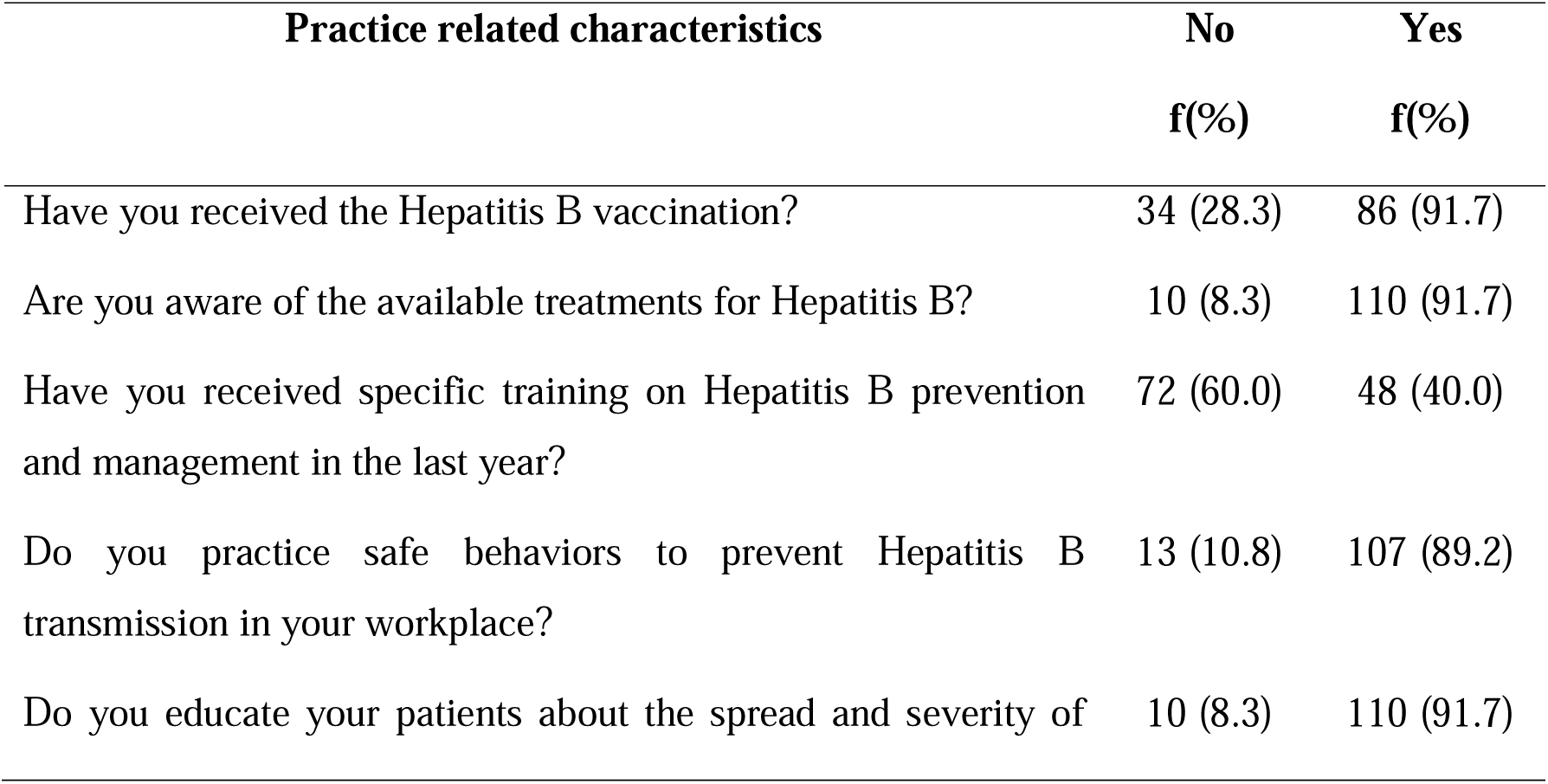
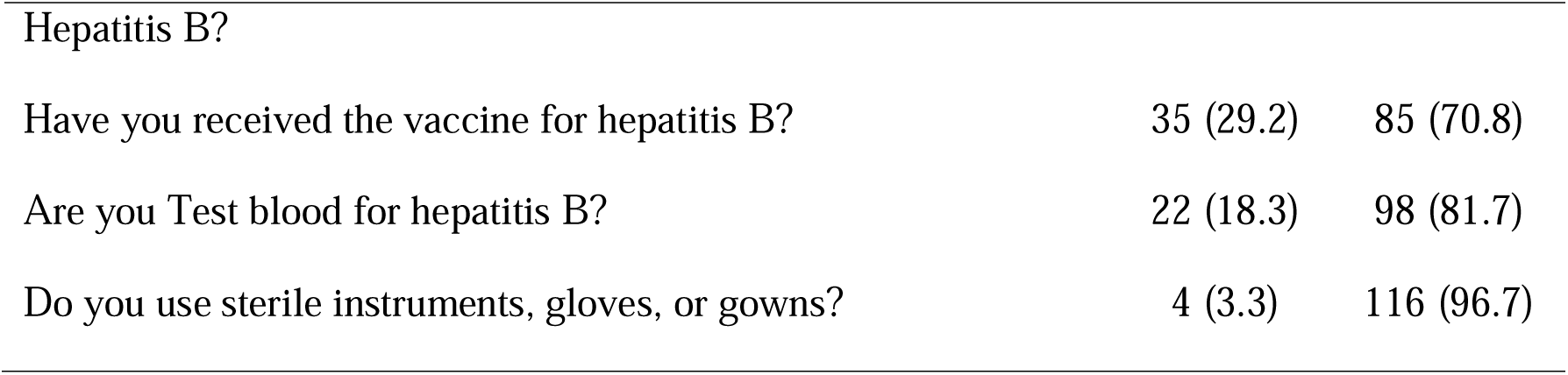
Distribution of practice regarding Hepatitis B Virus among Nurses.

| **Practice related characteristics** | **No**<br>**f(%)** | **Yes**<br>**f(%)** |
| --- | --- | --- |
| Have you received the Hepatitis B vaccination? | 34 (28.3) | 86 (91.7) |
| Are you aware of the available treatments for Hepatitis B? | 10 (8.3) | 110 (91.7) |
| Have you received specific training on Hepatitis B prevention and management in the last year? | 72 (60.0) | 48 (40.0) |
| Do you practice safe behaviors to prevent Hepatitis B transmission in your workplace? | 13 (10.8) | 107 (89.2) |
| Do you educate your patients about the spread and severity of | 10 (8.3) | 110 (91.7) |
| Hepatitis B? |  |  |
| Have you received the vaccine for hepatitis B? | 35 (29.2) | 85 (70.8) |
| Are you Test blood for hepatitis B? | 22 (18.3) | 98 (81.7) |
| Do you use sterile instruments, gloves, or gowns? | 4 (3.3) | 116 (96.7) |

### Overall knowledge and practice level of the respondents

Figure 1. depicts the level of average knowledge, attitude, and practices of respondents and their association with age. It can be seen that only 45.0% had average knowledge, while 25.0% had poor knowledge regarding HBV (Fig 1a). Similarly, only 43.3% of the respondents had a good attitude level while 28.3% had an average attitude level (Fig 1b). Additionally, 56.7% of nurses had good practice levels whereas 14.2% had average practice regarding HBV (Fig 1c). Approximately, over 48.3% of participants aged 41 to highest had a good attitude toward HBV, while the majority of respondents aged 31 to 40 years had an average attitude (Fig 1d).

**Fig 1.**
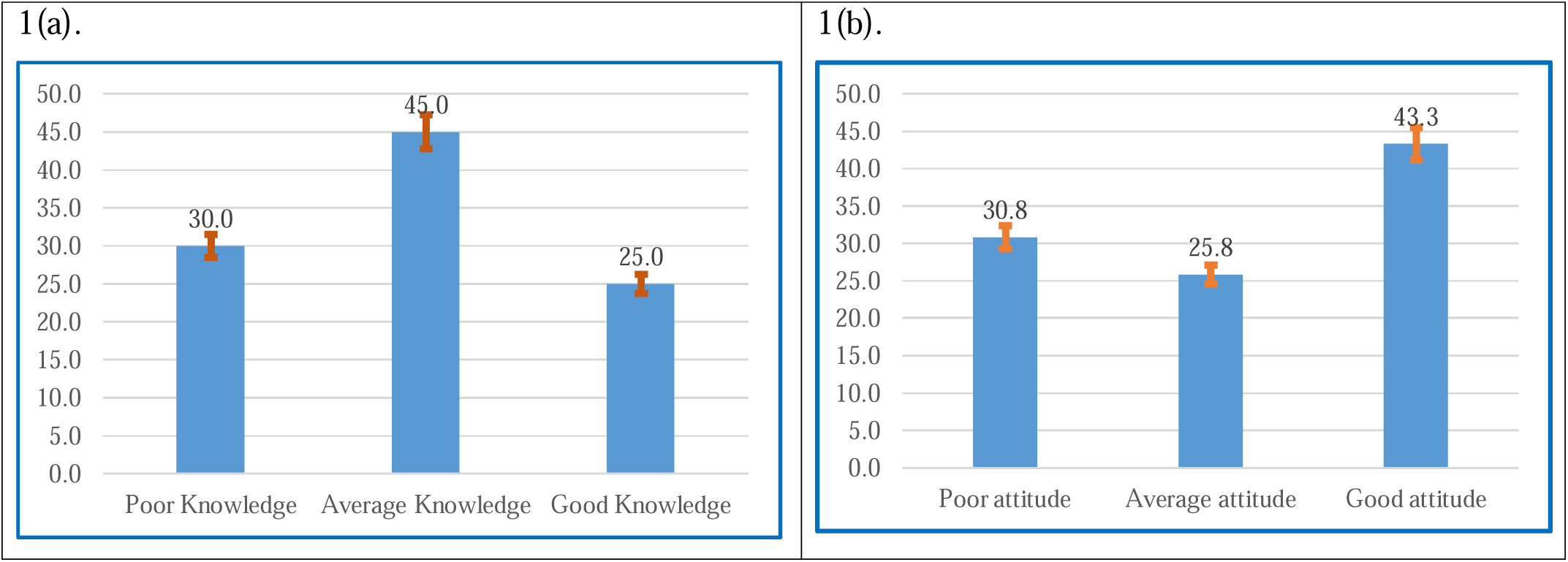
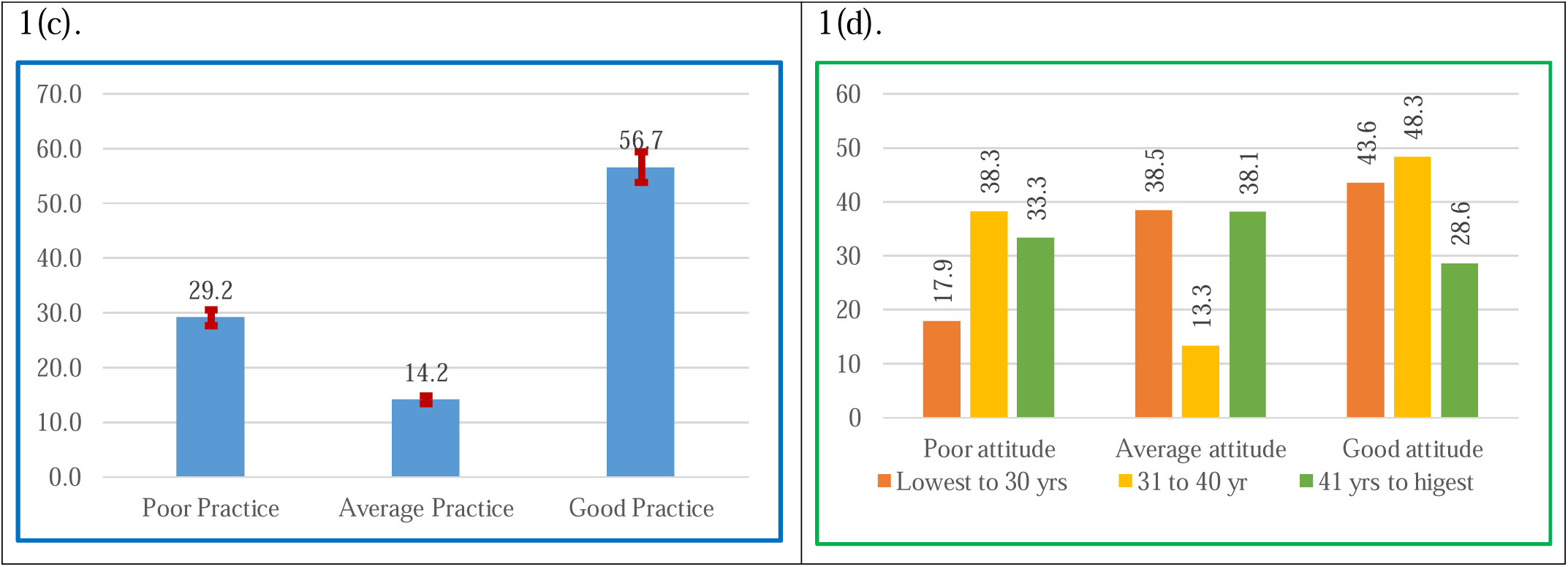
(a) Distribution of overall knowledge level; (b) Distribution of overall attitude level; (c) Distribution of overall practice level; (d) Association of attitude level with respondents’ age group.

### Variation in Knowledge, Attitude, and Practices of the respondents

A significant difference in respondents’ knowledge and practice with socio-demographic characteristics was observed (Table 5). Here, knowledge score was higher among rural participants [0.87 (0.81-0.92)versus 0.78 (0.77-0.79), p=0.001] than their counterparts. Similarly, the mean attitude score was significantly higher among the female participants compared to male participants: [mean attitude score: 0.88 (0.85-0.91) versus 0.77 (0.69-0.86), p=0.009]. Regarding practices, the mean practice score was associated with sex and religion. The practice score was significantly higher among female than male nurses [mean practice score: among female nurses [0.81 (0.77-0.85) versus male nurse 0.70 (0.59-0.81), p=0.02]. Similarly, the mean attitude score and practice score was significantly higher among non-muslim nurses than Muslim nurses: Attitude score: [0.87 (0.84-0.89) versus 0.63 (0.49-0.74), p=0.013] and practice score: [0.89 (0.76-0.93)versus 0.79 (0.61-0.87), p=0.035].

**Table 5.**
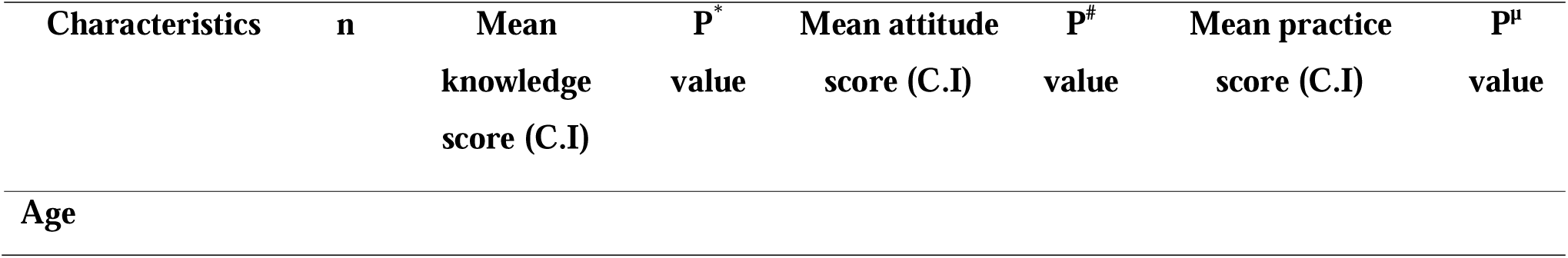
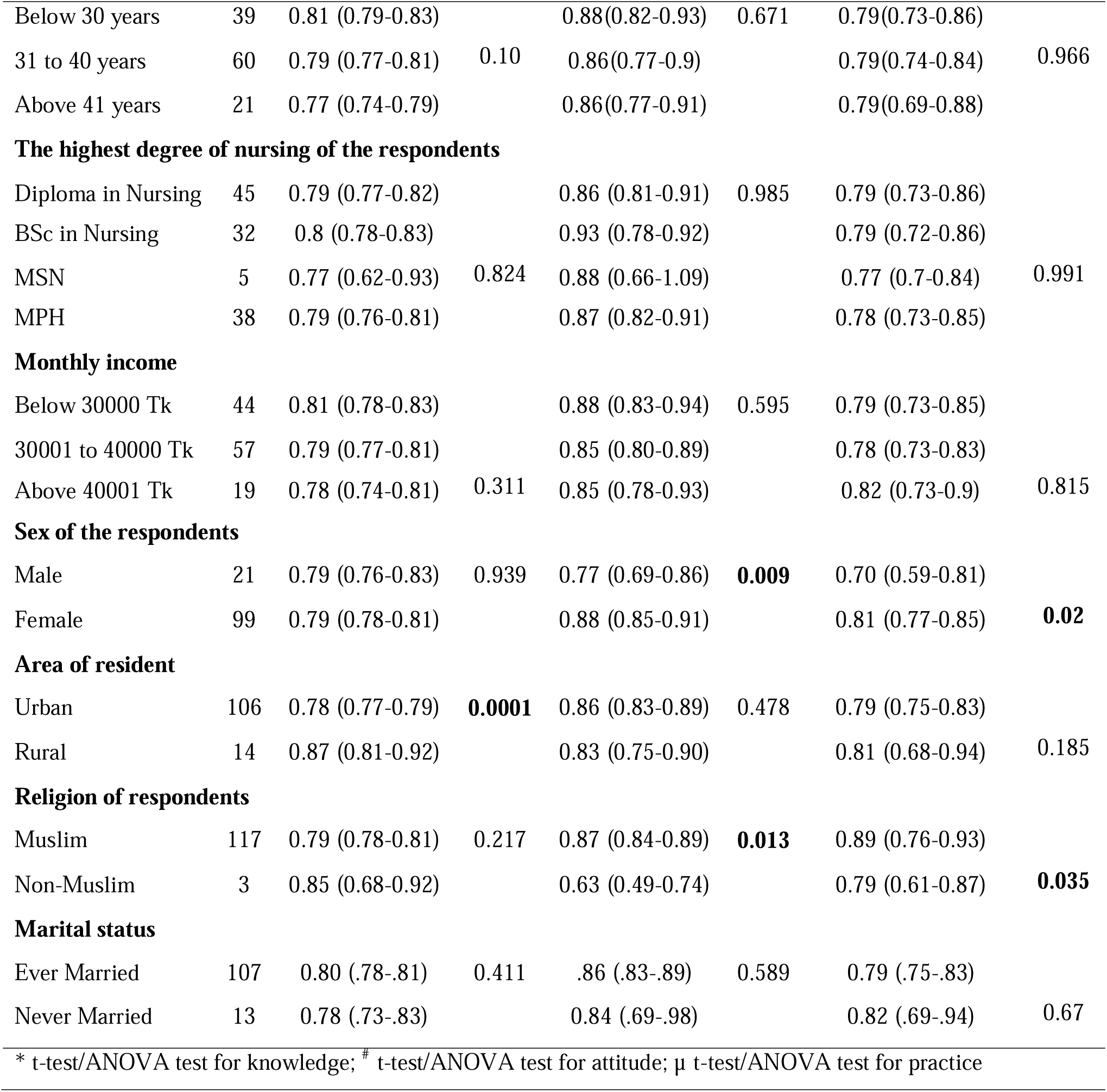
Independent sample t-test and one-way ANOVA test for knowledge, attitude, and practice score.

## Discussion

HCWs are at serious risk for occupational health hazards due to blood-borne pathogen exposure, especially HBV diseases. Due to their early exposure to hospital environments, healthcare workers, particularly nurses, are always at risk of contracting HBV from infected individuals [38]. To mitigate the risk of infection, it is crucial to possess accurate information, maintain a positive attitude toward prevention, and consistently adhere to hygienic practices. The sole aim of this study was to assess the KAP regarding HBV among nurses in Bangladesh.

The findings of this research indicate that 56.7% of nurses displayed proficient practice, 43.3% showed a favorable attitude, and 45.0% had a moderate understanding of HBV. The respondent’s area of residence had a notable correlation with their degree of knowledge. Age was shown to be associated with attitude, whereas gender and religion were strongly connected to activities related to HBV.

In the present study, almost half participants had average knowledge, and one-fourth of percent nurses had good knowledge regarding HBV. A similar study was conducted to assess Knowledge and preventive practices regarding HBV among nurses in some selected hospitals in Dhaka City, Bangladesh, and found that 30% of participants had poor knowledge regarding HBV [39]. In another study, The majority of the pre-clinical students (over 50%) were unaware of the symptoms, signs, and consequences of HBV infection [40].

Regarding attitude level, the current study found that the majority of the respondents had a good attitude regarding HBV. A study which was similar study conducted by Dwiartama et. Al., (2022) to assess KAP toward HBV Infection Prevention and Screening among Indonesians [41]. It found that 30.8% of respondents had a poor attitude about HBV, while the majority of the participants had a favorable attitude towards the preventive measures for HBV infection, which is very similar to this study. These results indicate that still there is a need for more HBV health promotion, targeted education, and training of nurses and midwives [42].

Regarding practice level, the majority of the participants had good practice and one-third of the respondents had poor practice towards the preventive measures for HBV infections. According to other studies, hepatitis is not well known among various populations, including healthcare workers, in several places across the world [43]. Though different results had been found in Sudan. A significant majority of healthcare workers, exceeding 80%, demonstrated awareness of the protocols for HBV. Furthermore, these professionals consistently adhered to practices such as sterilizing instruments, wearing gloves, etc [44], our study matched this result.

In the present investigation, it was discovered that there was a substantial connection between the level of knowledge and the location of the individual’s place of residence. In China, the knowledge level about HBV is connected to the area of residence. It was found that in the urban area respondents had good knowledge [45]. A similar result was found in Tanzania [46]. However, our study contradicts the previous studies. We found that the respondents from rural areas had a good knowledge. Another study conducted among HCWs in White Nile state in Sudan, on the contrary, presented that the level of knowledge was significantly associated with occupation and educational degree [42].

In Vietnam, a study about attitudes towards HBv concluded that there was not any significant relationship between the age of the respondents and attitude [47]. Our study revealed an opposite result. We found that age was connected with the attitude toward HBV which is consistent with a study in India. The Indian study found age as a significant factor in the positive attitude towards HBV [48]. Religion and marital status were not significantly associated with attitude of HBV infection. A similar study was conducted to assess the Prevalence of HBV and associated risk factors among adult patients at Dessie referral and Kemise general hospitals in northeastern Ethiopia [49].

It was shown that female HCWs in Afghanistan are more likely to demonstrate outstanding practices concerning HBV [50]. A different result was found in Saudi Arabia. They did not find any significant difference between the male and female HCWs regarding the practice of HBV [51]. Our study revealed that female nurses had more good practices regarding HBV. A lot of studies have shown that the job title and the highest nursing degree are major factors in determining KAP related to HBV [52]. However, our study did not find any statistically significant association between these features and KAP.

### Limitations and strengthens

A disadvantage of this research is the small sample size, as well as the fact that the respondents provided their self-reported data. The design of the research, which was cross-sectional, can also be considered one of the limitations. Despite the limitations, this study offers some important insights about KAP among the frontline HCWs—the nurses—about HBV for the policymakers and stakeholders. To the best of our knowledge, this is the first study conducted in Bangladesh among nurses regarding the KAP of HBV.

## Conclusion

In the study, the overall knowledge, attitude, and practice of nurses were not satisfactory, albeit the majority of them were able to prevent HBV. Furthermore, the respondent’s area of residency was significantly associated with knowledge level regarding HBV. Likewise, the respondent’s age was also significantly associated with an attitude toward HBV. The study participants had a low level of HBV vaccination coverage rate. Further strategies for preventing workplace exposure, training programs on HBV infection, including post-exposure prophylaxis (PEP), and increasing vaccination coverage rate of all HCWs, particularly nurses are highly recommended.

## Acknowledgments

We acknowledge the Institute of Biological Science, University of Rajshahi for their technical support during the study. We are also grateful to all participants included in this study.

## Ethical consent and permission for data collection

The permission of this study was approved by the Institutional Animal, Medical Ethics, Biosafety and Biosecuirity Committee (IAMEBBC) of Institute of Biological Sciences, University of Rajshahi [Ref. Memo No. 12(22)/320/IAMEBBC/IBSc]. Both written and verbal consent was taken from each participant before initiating the interview for data collection. A brief introduction on the aims and objectives of the study was given first and then, the written consent translated in Bengali. Participants who were agreed with the consent were finally included in the study.

## Authors contributions

**Conceptualizations-** Salekur Rahman, S M Shahinul Islam, Jahan Ara Khanam, and Mohammad Meshbahur Rahman

**Investigation**- Salekur Rahman, Sadhan Kumar Das, Zaki Farhana, Md Abu Bakkar Siddik, Anjan Kumar Roy, Piue Dey, Shuvojit Kumar Kundu, Md Anwar Hossain

**Methodology**- Salekur Rahman, Sadhan Kumar Das, Zaki Farhana, Md Abu Bakkar Siddik, Mohammad Meshbahur Rahman

**Supervision**- S M Shahinul Islam, Jahan Ara Khanam, and Mohammad Meshbahur Rahman

**Validation-** Salekur Rahman, Sadhan Kumar Das, Zaki Farhana, S M Shahinul Islam, Anton Abdulbasah Kamil, Jahan Ara Khanam, and Mohammad Meshbahur Rahman

**Visualization-** Salekur Rahman, Sadhan Kumar Das, Zaki Farhana, Anton Abdulbasah Kamil, Mohammad Meshbahur Rahman

**Fund acquisition-** Salekur Rahman, Sadhan Kumar Das, Zaki Farhana, Shuvojit Kumar Kundu, Anton Abdulbasah Kamil, and Mohammad Meshbahur Rahman

**Writing -review and editing-** Salekur Rahman, Sadhan Kumar Das, Zaki Farhana, Md Abu Bakkar Siddik, Anjan Kumar Roy, Piue Dey, Shuvojit Kumar Kundu, Md Anwar Hossain, S M Shahinul Islam, Anton Abdulbasah Kamil, Jahan Ara Khanam, and Mohammad Meshbahur Rahman

## Data availability statement

The datasets generated and/or analyzed during the current study are available from the corresponding author on reasonable request to.

